# PREDICT-ITB: Predicting response in children with dystonic CP to ITB – study protocol

**DOI:** 10.1101/2025.07.08.25331080

**Authors:** Sruthi P. Thomas, Darcy Fehlings, Sharon Ramey, Mark Conaway, Steven Kralik, Jeffrey Raskin

## Abstract

**Introduction:** Over 11,000 infants are diagnosed with cerebral palsy (CP) each year, with lifetime medical costs exceeding $1.4 million per person. Elevated muscle tone in CP, including dystonia and spasticity, significantly impairs function and quality of life. Intrathecal baclofen (ITB) is commonly used to manage dystonic CP, though evidence supporting its effectiveness is weak due to patient variability and study limitations. Spasticity may obscure dystonia symptoms, and factors like brain injury patterns and pain triggers are often overlooked in research. Despite uncertain effects on dystonia, ITB has shown benefits in pain relief, comfort, and caregiving ease. This proposal aims to evaluate ITB’s overall impact on children with dystonic CP, identify responders, and develop a comprehensive outcome measure using a prospective cohort study.

**Methods and Analysis:** We will conduct a prospective, observational study of 65 children with dystonia (Barry Albright Dystonia Scale (BADS) greater than 15) and CP who receive ITB. Changes in dystonia, spasticity, gross and fine motor function, and multiple patient-reported outcomes related to quality-of-life, depression, anxiety, pain and more. The primary analysis will use repeated measures models to estimate short and long-term changes from baseline in BADS scores at 3, 6 and 12-months. Secondary analysis will apply the same strategies to the other outcome measures. We will also conduct subgroup analysis and develop a multidimensional or composite measure.

**Ethics and dissemination:** Primary ethic approval was provided by the Baylor College of Medicine Institutional Review Board (H-54449). Results of the study will be disseminated via peer-reviewed presentations at scientific conferences and open access publication.

**Trial Registration Number:** NCT06606574 (clinicaltrials.gov)

**STRENGTHS AND LIMITATIONS:**

1. Unlike previous studies on ITB in dystonia and CP, a strength of this study is that it will directly measure effects of ITB beyond just dystonia, while also considering the child’s co- existing spasticity if present, known triggers of dystonia, including pain, and CNS injury patterns contributing to dystonia.
2. We consider multiple endpoints or the “total child” within the ICF Framework and whether concurrent therapeutic interventions appear to influence outcomes.
3. A limitation of the study is the lack of randomization to placebo or blinding.

## INTRODUCTION

Cerebral palsy (CP) is the most common cause of childhood-onset physical disability affecting ∼1 in 323 livebirths in the US (CDC). The lifetime medical costs for a child with CP are ∼$1.4 million.^1^ CP is defined as impairment of motor abilities due to non-progressive abnormal brain development or early life brain injury, with multiple etiologies and clinical presentations. One effective way to improve quality of life (QoL) and decrease disease burden is to significantly reduce or eliminate abnormally elevated muscle tone. When tone subsides, both active and passive movements are improved, in turn helping prevent musculoskeletal complications, improve quality of life, reduce pain, and ease the burden of daily care.

Dystonia, a devitalizing form of high muscle tone and movement disorder, affects >75% of children with CP and represents the predominant tone sub-type for an estimated 21%.^2^ Despite high prevalence, we have minimal evidence to guide treatment.^3–6^ Dystonia produces “involuntary sustained or intermittent muscle contractions [causing] twisting and repetitive movements, abnormal postures, or both”^7^ that cause pain/discomfort, disrupt sleep, create difficulty with seating, and impair daily caregiving tasks.”^8–11^ **Figure 1** captures the multiple serious effects of dystonia within an adaptation of the International Classification of Functioning, Disability, and Health (ICF) framework.

**Figure 1.**
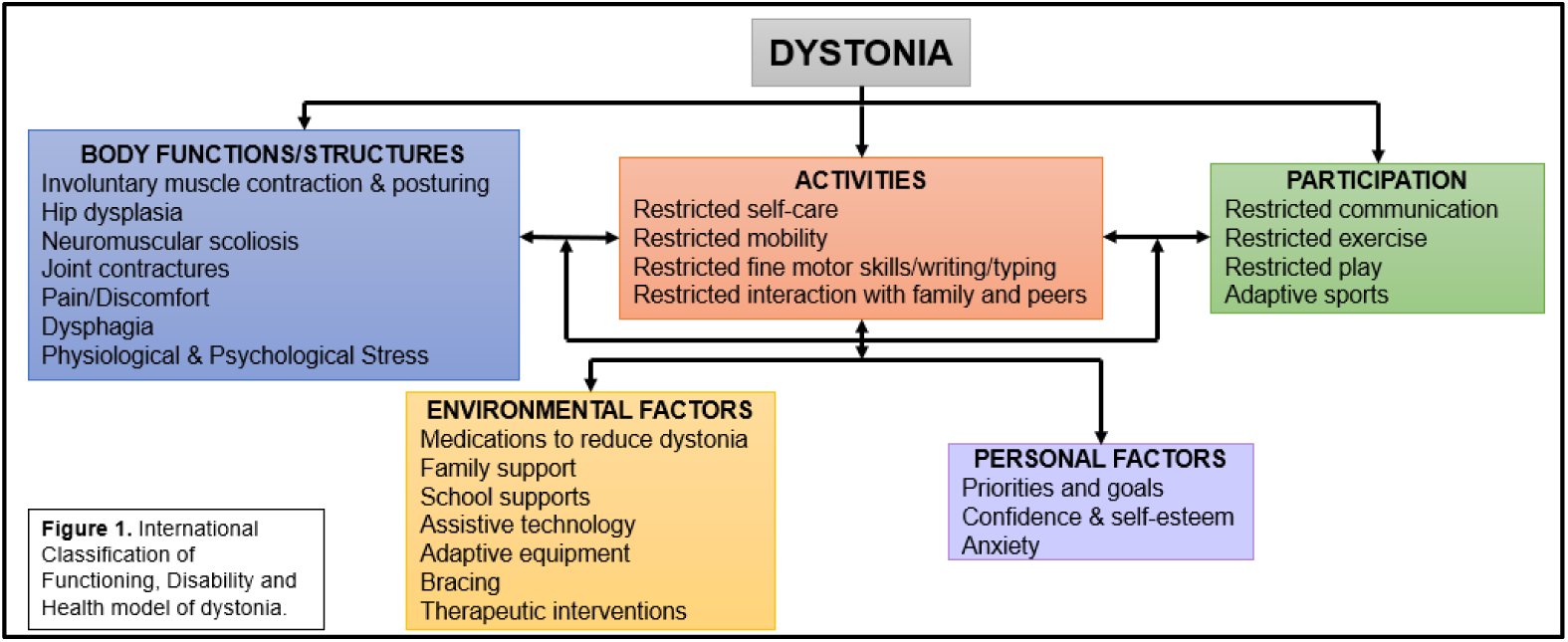
International Classification of Functioning, Disability and Health model dystonia.

For many children, dystonia co-exists with spasticity, a velocity-dependent tightness/stiffness of the muscles.^2^ The muscle overreacts to stimuli, such as rapid movement, resulting in stiffness and difficulty in voluntary motor control.^7^ In contrast to dystonia, there are many evidence-based FDA-approved therapies for spasticity, including enteral medications and more invasive neurosurgical approaches, notably intrathecal baclofen (ITB). Some treatments for spasticity overlap with those for dystonia, including ITB. There is a theoretical interplay between spasticity and dystonia, with spasticity masking dystonia by preventing muscular contractions, although this is poorly documented beyond clinical anecdote.^6,12^

Treatments applied, but not proven, to reduce dystonia include enteral drugs, injected neurotoxins and alcohols, positioning, ITB, and (rarely) deep brain stimulation (DBS). While ITB is FDA-approved for the reduction of spasticity, ITB has become the standard intervention for management of children with moderate-to-severe dystonic CP despite the lack of strong evidence of efficacy or effectiveness in reducing dystonia.^13,14^ ITB delivers baclofen that binds GABAβ receptors at the level of the spinal cord, reducing release of excitatory neurotransmitters and substance P, in turn reducing spasticity by blocking an overactive spinal reflex arc.^15–17^ However, it is unknown how this mechanism directly impacts dystonia. While baclofen can be taken enterally, delivery intrathecally via a threaded catheter connected to a motorized pump/reservoir implanted in the abdomen is significantly better.^18^ ITB was originally used for dystonia in a patient with refractory axial dystonia after scoliosis surgery, then later tested by Albright for dystonic CP.^19^ Beyond the high cost and burden of surgical pump/catheter implantation, ITB requires regular appointments for medication refills via transcutaneous pump access and dosage titration. The pump battery lasts 5-7 years then needs surgical replacement. If malfunction occurs, baclofen withdrawal can be severe, including death if untreated. Overdose causes significant central depression and potentially respiratory failure. Children require immediate assessment if either withdrawal or overdose risk arises.^20–22^ In summary, ITB is an invasive, expensive, and burdensome intervention despite its potential benefits. Accordingly, we need rigorous data about the degree to which ITB can reduce dystonia, identify characteristics of positive responders and assess its effect on motor function, pain, daily activities and participation, and QoL.^23^

A recent survey of CP experts in the U.S. and Canada documented wide variation in how clinicians view the use of ITB for dystonia. Some consistently rely on ITB to treat dystonia, while others remain skeptical and some choose alternatives such as DBS. Fehlings, Co-I for the proposed study, led a 2021 meta-analysis of interventions for dystonic CP; the team detected a small signal showing ITB reduced dystonia.^6^ Of concern, the only RCT in the meta-analysis showed little or no reduction in dystonia at the 3-month mark. However, it did show that ITB improved pain/comfort, sleep, ease of caregiving, and possibly motor function to varying degrees.^32^ **Table 1** provides a summary of key studies and identifies their weaknesses or limitations that are directly addressed in this revised proposal. None of these studies adequately addressed the following important issues: i) the presence or absence of co-morbid spasticity; ii) how ITB dosage titration progressed or if participants reached steady state before outcomes were measured (with the exception of the one RCT); iii) the concurrent use of orthotics, therapies, medications, and/or other surgical interventions overlapping with the study period; or iv) the relationship of CNS developmental abnormalities and injury patterns – known to vary considerably in dystonia - to treatment outcomes. The proposed study will directly measure effects of ITB beyond just dystonia, while also considering the child’s spasticity, known triggers of dystonia, including pain, and CNS injury patterns contributing to dystonia. Further, we consider multiple endpoints or the “total child” within the ICF Framework and whether concurrent therapeutic interventions appear to influence outcomes.

**TABLE 1.** Summary of studies of ITB’s effects on dystonia, including limits addressed by the proposed study. BADS = Barry Albright Dystonia Scale. QoL = quality of life. Burke-Fahn-Marsden Dystonia Rating Scale (BFMDRS), -M = Movement, -D = Disability. MA = Melbourne Assessment of unilateral upper limb function. GMFM = Gross Motor Function Measure. GAS = Goal Attainment Scale. MACS = Manual Ability Classification Scale. BFMF = Bimanual Fine Motor Function. DIS = Dyskinesia Impairment Scale. VAS = Visual Analogue Scale. PEDI = Pediatric Evaluation of Disability Inventory.

| **Reference** | **Design** | **Level of Evidence (for dystonia)** | **Participants** | **Primary Endpoints** | **Secondary Endpoints** | **Weaknesses & Limitations**<br>***ADDRESSED BY THIS PROPOSAL*** |
| --- | --- | --- | --- | --- | --- | --- |
| Albright et al., 1998<sup>19</sup> | Prospective case series with before/after evaluation | Class III | Generalized dystonic CP; age 4-42 years old (yo) (median 12) | BADS | None | -None had mixed spasticity + dystonia<br>-Only assessed dystonia<br>-Did not assess CNS injury pattern |
| Albright et al., 2001<sup>24</sup> | Prospective case series with before/after evaluation | Class III | Predominant severe dystonic CP; age 3-43 yo (median 13) | BADS | Patient & parent report-motor function, ease of care, QoL | -None had mixed spasticity + dystonia<br>-Did not assess CNS injury pattern<br>-Did not assess pain |
| Motta et al., 2008<sup>25</sup> | Prospective case series with before/after evaluation | Class III | Generalized dystonic CP; age 2-16 yo (mean 8) | BADS<br>BFMDR S-M | BFMDRS-D, Subjective report of motor function, | -None had mixed spasticity + dystonia |
|  |  |  |  |  | pain, ease of care | -Did not assess CNS injury pattern<br>-Did not assess Quality of Life (QoL) |
| Motta et al., 2009<sup>26</sup> | Prospective case series with before/after evaluation | Class III | Dystonic CP; age 11.3 ± 3.02 yo | BADS Total<br>BADS Upper Limb | MA Subjective evaluation of ease of care | -None had mixed spasticity + dystonia<br>-Did not assess CNS injury pattern<br>-Did not evaluate pain and QoL |
| Motta et al., 2011<sup>27</sup> | Prospective case series with before/after evaluation | Class III | Dystonic CP Mixed spastic-dystonic CP; age 15y11m ± 10y8m | BADS | GMFM | -Did not assess CNS injury pattern<br>-Did not evaluation pain, ease of care, or QoL<br>-Did not account for effects of spasticity |
| Eek et al., 2018<sup>28</sup> | Retrospective case series with before/active evaluation | Class III | Dyskinetic CP with dystonia; Age 10y11m ± 4y9m | BADS | GAS, GMFM-88, MACS, BFMF. Parent rating of PAIN, motor function, ease of care | -None had mixed spasticity + dystonia<br>-Retrospective data only<br>-Did not assess CNS injury pattern<br>-Did not evaluate QoL |
| Kim et al., 2019<sup>29</sup> | Retrospective case series with before/active evaluation | Class III | Mixed tone; Age 37.70 ± 7.02 yo | BFMDR S-M | BFMDRS-D Visual Acuity Scale SF-36 | -No children, all adults<br>-Did not assess CNS injury pattern<br>-Did not account for effects of spasticity |
| Bonouverie et al., 2019<sup>30</sup> | Randomized double-blinded placebo-controlled trial | Class II | Non-ambulatory dyskinetic CP; Age 7-24.4 yo (mean 16) | GAS | BADS DIS | -Patients only on ITB for 3 months – possibly did not reach ideal dosing<br>-Primary outcome was GAS – potentially<br>**underpowered for dystonia outcomes**<br>**-Did not assess CNS injury pattern**<br>**-Did not account for effects of spasticity** |
| Bonouverié et al., 2022 (follow-up to 2019 RCT)<sup>3</sup><br>1 | Prospective cohort study | Class III | Non-ambulatory dyskinetic CP; Age 7-24.4 yo (mean 16) | GAS | BADS DIS | **-Unable to separate placebo from true effects for patient reported outcomes**<br>**-Did not consider titration after 3 months**<br>**-Did not account for effects of spasticity** |
| Bonouverié et al., 2023 (follow-up to 2019 RCT)<sup>3</sup><br>2 | Prospective cohort study | Class III | Non-ambulatory dyskinetic CP; Age 7-24.4 yo (mean 16) | GAS | BADS, DIS. Passive ROM, Spasticity H/M ratio of soleus, Pain with VAS PEDI | **-Unable to separate placebo from true effect for patient reported outcomes**<br>**-Characterized brain injury patterns but not analyzed in relation to outcomes**<br>**-Did not account for effects of spasticity** |

Clinical anecdote and published commentary hypothesize that spasticity reduction may unmask dystonia.^6,12^ By reducing the tonic stretch reflex associated with spasticity, muscles contract with more force, potentially amplifying dystonia. To our knowledge, only one study investigated spasticity reduction unmasking dystonia, reporting a correlation between unmasking and congenital/genetic conditions when children underwent selective dorsal rhizotomy, a neurosurgical intervention to reduce spasticity.^33^ Thomas (PI) published findings that children with spasticity alone required lower ITB dosage and those with dystonia required higher dosage.^34^ Theoretically, when ITB starts, spasticity will be lowered but dystonia could remain the same or initially worsen with unmasking. Either mechanism could improve outcomes. We hypothesize that children with co-morbid spasticity will see significantly higher magnitude benefits from ITB than do those who are predominantly dystonic, regardless of the effect on dystonia.

Certain stimuli can trigger dystonia of all etiologies, including pain, anxiety, stress, heightened emotions, and fatigue.^35^ Further, dystonia can be quite painful, leading to unclear directionality of correlations.^36^ Baclofen has been shown to reduce substance P release in the spinal cord in animal models.^17^ Substance P increases pain perception by altering cellular signaling pathways at the level of the dorsal horn in the spinal cord.^37^ We hypothesize that children with dystonic CP and concurrent high pain will experience greater dystonia reduction as well as improved QoL after ITB initiation.

Dystonia is thought to result from malformation of or injury to the basal ganglia, cerebellum, cortex, thalamus, and brainstem.^38^ Recently, injury to the internal capsule/corona radiata (periventricular leukomalacia (PVL)) has also been identified as a potential cause of dystonia, important because this type of injury occurs with high prevalence in CP.^39^ Previously, PVL was only associated with spastic CP.^40^ Several studies have sought to characterize brain injury patterns. Most relied on focused analysis, with a priori identification of areas of interest.^41,42^ To our knowledge, no whole brain analyses or tract-based spatial statistics have measured white matter tract injuries in this population. Additionally, no reports have related CNS injury patterns to tone management.^43^ We hypothesize that children with dystonic CP resulting from white matter injury, such as PVL, will show an overall significantly higher positive response to ITB than those with dystonia resulting from deeper grey matter changes.

The primary objective of this study is to determine if ITB reduces dystonia as measured by the Barry Albright Dystonia Scale (BADS).^44^ In addition to changes in quantitative dystonia outcomes, this study will investigate if ITB affects the functional impact of dystonia, spasticity, function, pain, and other patient-reported outcomes (PROs), including quality of life, anxiety, and depression. This detailed characterization will be used to look for patient characteristics associated with positive outcome(s), including variation based on initial quantity of spasticity and baseline pain. Further, this broad set of outcome measures will allow us to identify which outcome measures are most sensitive to capturing changes post-ITB and these will be used to create a multidimensional or composite measure for future ITB studies and for clinical use. A multidimensional or composite measure will save time and resources while reducing the burden on patients and their families in future studies by narrowing the battery of measured outcomes.

Another secondary objective is to use this patient population to tease apart more of the pathophysiology of secondary dystonia in CP. This study will complete a detailed characterization of brain malformation and injury patterns of the children enrolled in this study. This will include four types of analysis using MRI brain imaging: 1) Classification according to the MRI Classification Scale for CP (MRICS), 2) Whole brain analysis to calculate whole brain grey and white matter volumes, regional volumes, and cortical thickness, 3) Lesion network mapping – areas of abnormal anatomy will be mapped on to known pediatric connectivity maps, and 4) Tract-based spatial statistics (TBSS) – quantitative analysis of white matter tracts.

## METHODS AND ANALYSIS

### Study Design

We will recruit 65 children over 40 months (3.25 years, March 2025-June 2028) to be in our study. Outcome assessments will be at baseline and 3, 6, and 12 months post-ITB implantation. Recruitment materials will include dual language (English, Spanish) family-friendly print and video materials and web-based study information. The study Parent/Patient Advisory Council will review all recruitment materials to ensure easy comprehension and maximize addressing questions of high importance to families/patients. A prospective longitudinal cohort design was chosen for multiple reasons. First, it allows for the collection of a rich, comprehensive set of data. Second, it allows us to track outcomes in real-world practice settings. Third, many of these children are seeking ITB because of the moderate-to-severe tone that they and their medical providers feel are contributing to significant comorbidity. As a result, this posed ethical challenges to having a placebo control.

### Recruitment, inclusion, and exclusion criteria

This prospective longitudinal cohort study will be conducted at Texas Children’s Hospital in Houston, TX. Patients will be screened for eligibility once they have decided to proceed with pump placement.

#### Inclusion criteria

Child must be large enough to have an ITB pump implanted (typically ∼18 kg/4 years old); <18 yrs old; diagnosis of CP with dystonia verified via Hypertonia Assessment Tool (HAT) and Barry-Albright Dystonia Scale (BADS) score >14; identified by a physician as a candidate for ITB treatment for tone management; and the family/child have agreed to proceed with implantation.^45^

#### Exclusion criteria

Urgent need for ITB such as status dystonicus or paroxysmal sympathetic hyperactivity; Botulinum injections within past 3 months or phenol injections within past 6 months; foster care placement or incarceration; family not willing to participate in full schedule of assessments and 12-month follow-up.

### Sample size calculation

Power was assessed for the primary outcome - specifically, a significant improvement at 12- months post-intervention on the BADS score. Combining the results from multiple studies, we estimate the standard deviation (σ^) of pre-intervention/baseline and 12-month BADS scores at 4.69.^19,24,27,28,30,46,47^ The mean pre-intervention BADS score was 19.7 and changes at 12- months ranged from 15% to 50% improvement. Of note, the minimal clinically important difference (MCID) for the BADS is a change of 25% based on observational ITB studies by Albright.^19,48^ Using a conservative estimate of zero correlation between baseline and 12-month BADS scores, the σ of changes from baseline = 6.69 (√2σ^). With a final sample of 55 children, the paired t-test has 90% power, with a 2-sided significance level of 5%, for a change of 15%. Allowing for a 15% dropout, the required initial recruitment sample size is 65 children. Despite relatively high complication rates noted in publications, the one RCT in this field kept patients enrolled with only short-term complications that did not lead to study withdrawal.^31^ For pumps implanted between 4/1/21-3/31/22 (1 year), the attrition rate at TCH was only 4% (n=28), well below our conservative estimate of 15% for the sample size calculation. We aim to enroll 15 children in Year 1, 20 children/yr in both Years 2 & 3, and 10 children in Year 4. Based on a review of current patients with ITB, 44% would meet enrollment criteria. We implanted 32 pumps in 2021, 35 in 2022, 27 in 2023, and 12 in 2024. We have implanted 8 so far in 2025. Thus, our enrollment feasibility is strongly supported. We propose data analyses through 12- month follow-up using repeated measures models, accounting for factors such as age, sex, and baseline severity of dystonia and/or spasticity, using the F-test within these models to test for significant changes in 12-month outcomes, controlling for multiple tests/comparisons. Inclusion of significant predictors will increase the power and precision for estimating changes, as does a positive within-subject correlation between baseline and 12-month scores. The use of repeated measures models also increases the power since inclusion of data from subjects up to the point of dropout increases the degrees of freedom for error.

### Blinding procedures

Since this is a prospective observational study, children and their families are not blinded to the intervention. Therapists completing functional assessments will not be aware of the child’s current ITB dose. Video review to measure BADS and DIS-II will be completed in unlabeled batches to minimize bias.

### Study intervention

#### Intrathecal baclofen

All children will receive a Medtronic SynchroMed^TM^ III pump with Ascenda^TM^ catheter, the most used pump/catheter brand for children in the U.S. Catheter tip positioning will be co-determined by the primary PM&R physician and implanting neurosurgeon. The bulk of children with dystonia on the BADS >14 typically have involvement of all limbs ± trunk, in which case the catheter tip would be placed in the cervical region. While we will be rapidly up-titrating baclofen, we will start with 500 mcg/ml concentration in case the child is highly sensitive to ITB and needs to be down-titrated below 100 mcg/day due to initial side effects, e.g., urinary retention, somnolence. This allows us to drop as low as 24 mcg/day. We will use Lioresal® brand baclofen for ITB, adding more consistency to the protocol. All children will be converted to 2000 mcg/ml concentration if their total daily dose is >200 mcg/day.

#### Dose titration schedule & Rationale for dosing strategy

The titration algorithm for ITB shown in **Figure 2** parallels the titration protocol used by the IDYS trial, the only RCT study of ITB and dystonic CP.^49^ This algorithm’s titration rate is backed by the protocol described by Saulino’s group, a protocol that has become standard in ITB programs and parallels typical titration plans.^50^ Twelve months of receiving treatment will be highly informative in assessing sustainability of improvements (if these occur) and allow most children to reach a steady state with their dosing. We reviewed all ITB implantations at TCH between 4/1/21-3/31/22 (n=28) to compare current practices to those proposed in the ITB titration protocol. The average ITB dose at the time of hospital discharge post-implantation was 172 mcg/day (65-375 mcg/day) and the average current dose is 417 mcg/day (72-904 mcg, ≥12 months post-ITB initiation). The absolute average titration percentage at outpatient visits was 12% (5-20%). The proposed ITB titration plan titrates by 10-20% based on the current ITB dose, which is in line with this data.

**Figure 2.**
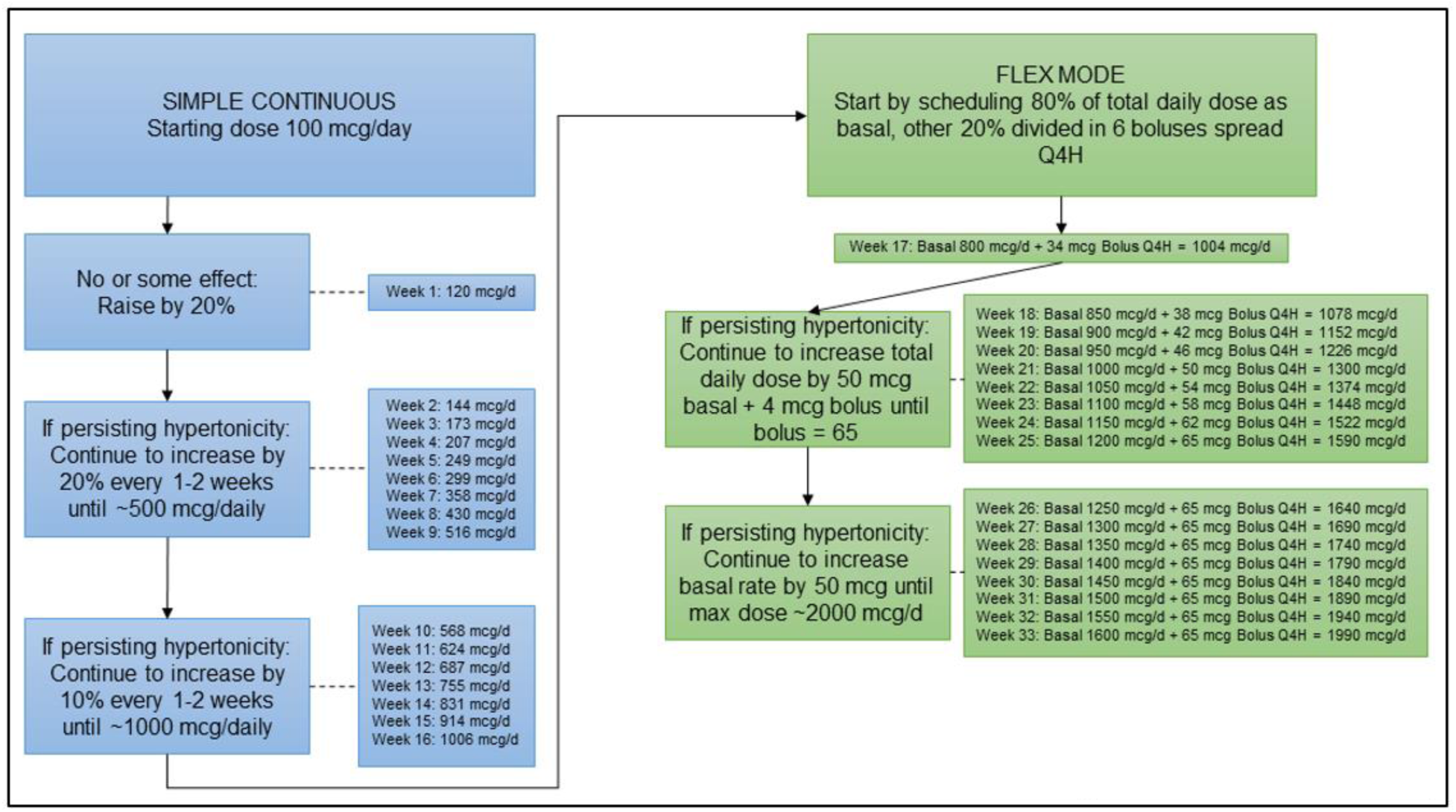
ITB Titration Protocal.

#### Dose modification strategy

The ITB dose will be *<u>increased</u>* to the next step if 1) there is room for improvement on the Dyskinetic CP Functional Impact Scale (D-FIS) and/or persistent spasticity based on Australian Spasticity Assessment Scale (ASAS) total, and 2) there are no detrimental side effects.^51^ This will continue until scores are steady for 2 consecutive visits. The ITB dose will be *<u>decreased</u>* to the previous step if 1) scores have been steady for 2 visits or 2) there are detrimental side effects. Once steady state is reached, this new dose becomes the <u>maintenance dose</u> for the rest of treatment. If detrimental side effects occur, we will see the child weekly to assist with and confirm resolution of side effects. If a child is very sensitive to titrations, meaning there is significant sedation or urinary retention, the frequency of titration can be slowed as much as necessary.

### Data collection and data management

All data will be collected in a HIPAA-compliant REDCap database with subjects deidentified. The research team will have access to the REDCap database and will also be able to download data from REDCap as .xlsx files for analyses. Raw MRI data will not have to leave the Texas Children’s Hospital system, as the pediatric neuroradiologist completing the analyses is also located here. Dr. Kralik will take image sequences from the local Picture Archiving and Communication System (PACS) and transfer them on to a local computer for tract-based spatial statistics. Diffusion tensor imaging sequences from the MRI brain in this study will be analyzed using whole brain analysis. Quantitative analysis will be performed using the volumetric 3D T1- weighted sequence using Freesurfer software (Version 5.3.0), which is publicly available. Post-analysis data will be entered into REDCap. The Office of Sponsored Programs at Baylor College of Medicine that is administering this award has created a data management and sharing plan compliance system as part of their process for submitting the annual NIH progress report. The Office will collect information related to the number of research participants that are deposited each reporting year.

Given the complex nature of this intervention protocol, all clinicians in our ITB clinic will be trained on the study design, use of the titration protocol, the side effects questionnaire, D-FIS, and ASAS. Annual refreshers will ensure high quality compliance and minimize any study drift. We will use an Epic flowsheet to simplify tracking and compliance with data collection. Thomas (PI) will conduct quarterly checks of all enrolled and treated children to confirm that the ITB titration protocol is being administered and recorded accurately. Any discrepancies will be tracked and reported to the Data Safety Monitoring Committee (DSMC) and corrective actions will be taken, if needed.

### Outcomes

#### Primary outcome

Barry-Albright Dystonia Scale (BADS). The BADS is based on the Burke-Fahn-Marsden Dystonia Rating Scale, originally designed to detect cervical dystonia in adults.^52^ The BADS tailors this scale for detecting dystonia in CP and acquired brain injury in individuals with impaired cognition and physical ability. Scoring focuses on posturing and involuntary dystonic movements to detect potential changes in dystonia that affect ease of care and comfort. It does not assess functional tasks. It has moderate internal consistency and interrater reliability. It has effectively detected change with ITB.^27,46,53–55^ The BADS was also found to have the greatest clinical utility for children with dystonic CP.^56^

#### Secondary outcomes

Dyskinesia Impairment Scale-II (DIS-II). The DIS-II is one of the newest scales to measure dystonia and choreoathetosis at rest and during activity.^57^ It is unique in separating choreoathetosis from dystonia in this population. It has moderate internal consistency and fair interrater reliability.^58,59^ We also include this measure because it was a key measure in the IDYS trial by Bonouvrie LA *et al.*, the single RCT trial, thus permitting direct comparison across our studies.^30^

D-FIS.^51^ Dystonia is understood by clinicians and described by caregivers as follows: every time someone tries to move or is handled by someone else, a part of their body moves out of their control.^60^ Therefore, even “mild” dystonia by some assessment scales can have significant functional impact. Similarly, even “small” decreases in dystonia by some assessment scales can yield significant functional improvement. The ideal scale to assess the real-world impact of dystonia on children with CP is a scale that assesses the functional impact experienced by the child with dystonia. This scale is the D-FIS. The D-FIS was created in partnership with 8 primary caregivers of children with dyskinetic CP and guidance from occupational, physical, and speech therapists, pediatric neurologists, and pediatric physiatrists. Participants (children with CP) and their caregivers report how much dystonia interferes with different aspects of their life. The D- FIS captures meaningful change in dystonia, which other quantitative dystonia measures could miss. Though the D-FIS is a new scale, it has been cross-validated against the Barry Albright Dystonia Scale (BADS), one of the most used scales for dystonia treatment assessment in CP and the longest standing scale in existence for assessment of dystonia in people with CP.

Australian Spasticity Assessment Scale (ASAS) and Composite Measure. It has clear instructions on how the limb must be tested while supine to increase reproducibility. Its interrater reliability is very high with a weighted kappa of 0.87, making it ideal for a large study with multiple time points.^61^ We will generate a composite score (as well have individual scores) for key muscle groups (**Table 3**)

**Table 3.** Composite ASAS Score. Score range 0-48. If uppers are analyzed separately from lowers, scale 0-24 per section.

| Left | Upper Limbs | Right |
| --- | --- | --- |
| 0-4 | Shoulder Adductors | 0-4 |
| 0-4 | Elbow flexors | 0-4 |
| 0-4 | Wrist flexors | 0-4 |
|  | Lower Limbs |  |
| 0-4 | Hip adductors with knees flexed | 0-4 |
| 0-4 | Knee flexors | 0-4 |
| 0-4 | Ankle plantarflexors with knees extended | 0-4 |

Modified Ashworth Scale (MAS). While our study relies strongly on the ASAS, we will also measure MAS so our data can be compared across studies.^62^

Gross Motor Function Measure (GMFM)-88. The GMFM-88 is an assessment tool to measure changes in gross motor function with interventions in children with CP. Given that children enrolled in this study will fall in the GMFCS IV-V category, we will not score section E: walking, running, and jumping. As a widely used clinical and research tool, it has an excellent test-retest reliability and excellent inter- and intrarater reliability.^63,64^ The minimal clinically important difference (MCID) is ∼0.7-1.2 points in ambulatory children with CP but unfortunately a level has not been determined for non-ambulatory children.^65^

Box and Blocks Test. This test is a quick, simple way to assess manual dexterity. This test cannot be used in a child with severe upper extremity impairment or severe cognitive impairment, therefore, not all participants in this study will be able to complete this task. A child is asked to move as many blocks as possible from one side of a divided box to the other in 60 seconds. It has excellent interrater reliability and MCID of 5.29-6.46 seconds.^66,67^

Performance Quality Rating Scale (PQRS). The PQRS is an observational measure of performance quality of a parent/patient-selected meaningful activity. Occupational therapists use a video recording of a child performing the task of interest and then rate the quality. The task is chosen after discussion between the family, occupational therapist, and child if possible. It has high interrater reliability from 0.83-0.93 and test-retest reliability >0.80.^68^

Canadian Occupational Performance Measure (COPM). The COPM focuses on occupational performance in all areas of life, including self-care, leisure, and productivity. It consists of a semi-structured interview to determine areas of concern for the parent and child. Together with an occupational therapist, they set important goals and then score performance on the particular concerns at the set timepoints in the study. We will have families and children set 2-3 goals that are relevant to ITB treatment. The COPM is an excellent tool for this study as it is very sensitive to change and has an established MCID of 2 points.^69^

Caregiver Priorities & Child Health Index of Life with Disabilities (CPCHILD^©^). The CPCHILD^©^ measures a child with severe disability’s overall health, comfort, function, and QoL in relationship to their medical conditions. It has 37 questions answered by the child or parent about activities of daily living, positioning, transferring and mobility, comfort and emotions, communications and social interaction, health, and overall QoL. It was initially validated in children with CP and traumatic brain injuries. Prospective longitudinal cohort studies are underway to assess the measure’s responsiveness/sensitivity to change.^70^

Patient-Reported Outcomes Measurement Information System (PROMIS^®^) Questionnaires. NIH’s Office of Strategic Coordination – Common Fund’s PROMIS^®^ developed a validated item bank of patient-reported outcomes for clinical research and practice. The PROMIS offers many short form questionnaires that are simple and quick for parents and children to complete. We will use the following to assess pain, anxiety, depression, and stress:

PROMIS^®^ Pediatric Item Bank v1.0 – Pain Behavior 8a/PROMIS^®^ Parent Proxy Bank v1.0 –Pain Behavior 8a.

PROMIS^®^ Pediatric Item Bank v2.0 – Pediatric Anxiety – Short Form 8a/ Parent Proxy Bank v2.0 – Parent Proxy Anxiety – Short Form 8a.

PROMIS^®^ Pediatric Item Bank v2.0 – Pediatric Depressive Symptoms – Short Form 8a/ Parent Proxy Bank v2.0 – Parent Proxy Depressive Symptoms – Short Form 6a.

PROMIS^®^ Parent Proxy Bank v1.0 – Psychological Stress Experiences – Short Form 4a. PROMIS^®^ Parent Proxy Bank v1.0 – Physical Stress Experiences – Short Form 4a.

Goal Attainment Scale (GAS). It selects a single important goal to be tracked before, during, and after an intervention. GAS was created to account for the heterogeneity of the rehabilitation population. In addition to choosing the goal(s), the child and their family, with guidance from their care team, choose how to define a successful outcome. The expected outcome will be defined as 0. Better than expected performance will be defined as +1 or +2. Poorer than expected performance will be defined as -1 or -2. GAS is tailored to each child and has been shown to be as sensitive to change over time as other standard outcome measurement tools, including the GMFM (discussed above) and the Pediatric Evaluation of Disability Inventory.^71^ The single RCT IDYS trial used the GAS as their 1° outcome measure. By including this, we can directly compare results.^30^

#### Additional captured data

##### Demographics & Concurrent Management

We will use an intake questionnaire to collect pertinent demographic and clinical history information, including birthdate, sex, race/ethnicity, home zip code, parental education, household income, age at implantation, etiology and severity indicators of the child’s CP, and surgical history. We will obtain an inventory of the child’s use of orthotics, therapies, and prescribed medications, as well as any complementary or alternative approaches (e.g., massage, acupuncture, chiropractic practices). *We will also track the position of each child’s ITB catheter tip, as variability in the catheter placement could possibly affect outcomes.* Some information will be gathered from sections 5-9 of the *Caregiver Priorities & Child Health Index of Life with Disabilities (CPCHILD^©^).* There will also be a separate brief questionnaire that asks families to update investigators with any changes in orthotics, therapies, or other adjunctive interventions.

##### Neuroimaging

MRI Brain imaging of all children enrolled in the study will be accessed on the local PACS system for deeper analysis. The original neuroradiology imaging impression will be reviewed. If the patient does not have a MRI brain available from before ITB implant, research funds can be used to acquire one if there is no clinical reason to obtain it. Patients and their families may opt out of the MRI brain and remain in the study.

### Statistical analysis

#### Primary outcome

The primary analyses for Aim 1 will use repeated measures models, carried out in SAS PROC MIXED, to estimate short and long-term changes from baseline in BADS scores at 3, 6 and 12- months. We anticipate the use of a spatial-power covariance matrix, but will also consider other covariance structures, such as mixed models. The model will include terms for baseline dystonia, spasticity, and pain; F-tests will be used to test the significance of these predictors. To assess if trajectories of ITB changes differ by predictors, interactions between these and assessment time will be added to the model.

#### Secondary outcomes

##### Additional endpoints

Analyses similar to those for Aim 1 will be conducted for the DIS, D-FIS, ASAS, MAS, GMFM-66, Box and Blocks Test, PQRS, COPM and PROs. Within each specific set of analyses, a false discovery rate of 5% will be used to adjust for testing across multiple secondary outcomes.

##### Subgroup analysis

Changes in specific subpopulations and comparing differential patterns of response to ITB in subpopulations will be accomplished by adding main effects and interactions to this model. Specifically, to assess the effect of ITB within the mixed tone population (Aim 1), a main effect for mixed tone (yes/no) and a mixed tone by assessment time interaction term will be added to the model. Contrasts within the model will be used to estimate the effect of ITB on changes over time within the mixed tone subpopulation and then be compared with changes that occur in the primarily dystonic subpopulation. Similar exploratory analyses will be done to compare response to ITB in the low and high spasticity subpopulations and those with significant baseline pain. Post hoc analyses will consider whether concurrent treatments and other variables possibly contributed to different outcome patterns during ITB treatment and at 12-months for these subpopulations.

##### Development of a multidimensional or composite measure

We will implement several analytic strategies that include multiple endpoints. One example is O’Brien’s method that scores each child’s rank order of change (gain, improvement) on a selected set of outcome measures and then tests whether certain subpopulations have significant differences in their mean magnitude of change from baseline.^72^ This method does not, however, provide information about the types of changes that occur. Other methods ascribe differential weight to the outcome measures, perhaps related to the child’s baseline clinical profile. While not originally intended for this, O’Brien’s method can also be used to cluster children into, for example, low, medium, or high changes from baseline, as measured by their average rank. These classifications are relative to the children in the sample, since children with ‘low’ average ranks may have responded well to therapy but not as well as others in the sample. Subsequently, a cluster analysis will be applied to changes at 12 months to identify groups of children who have similar content changes (e.g., spasticity, dystonia, pain, QoL) within the battery of outcomes. Initially we will use k-means clustering, starting with 3 clusters, although we will consider other numbers of clusters as guided by the gap statistic. The k-means clustering differs from the use of the average rank in that the clusters identify children who respond similarly, which could mean large changes on some outcomes and smaller changes on others. These initial analyses will be followed by other methods of clustering, including hierarchical methods. Multivariate regression strategies will be used to explore which characteristics of participants differ among the response/outcome clusters.^73^

##### Assess representation of the multidimensional or composite measure

Based on initial analyses described above, various dimension reduction methods will be used to explore whether a subset of the outcome measures adequately represents significant changes across the full set of outcome measures. The analyses will begin simply by computing correlations among changes in outcome variables and identifying patterns of outcomes that are highly related to each other. Subsequent analyses will use principal components analyses and multidimensional scaling.^74^

##### Preliminary exploration of the multidimensional or composite endpoint

For these analyses, we use the term ‘composite endpoint’ to mean a single endpoint constructed from parts or all available endpoints. Conaway (Statistician for our study and Co-I) led a team that developed the ‘Myasthenia Gravis Composite’ (MGC) in which individual items were selected from existing MG-specific scales “so as to be meaningful to both the physician and the patient, frequently abnormal in patients with active disease, and responsive to clinical change.”^75^ We are creating a similar composite tool for children with dystonic CP. The MGC relied heavily on clinician expertise in choosing the initial set of items, which were then tested and modified based on the performance in two clinical trials. In the current proposal, based on the clinical expertise of the research team and our Parent/Patient Council, we will select an initial set of items and evaluate the responsiveness of these items to clinical change, refining the items as needed. Subsequent inquiry will further refine the items selected and establish psychometric properties and sensitivity/specificity of the new composite tool.

##### Neuroimaging analysis

Children will complete non-contrast brain MRIs prior to placement of the baclofen pump – which is standard clinical practice. All MRI scans will be performed on a 3 Tesla MRI scanner (Siemens, Erlangen Germany) at Texas Children’s Hospital. MRI sequences will include volumetric 3D T1W, T2W, susceptibility-weighted imaging (SWI), FLAIR, and diffusion tensor imaging (DTI) with 32 directions. Stephen Kralik (Co-I) with 10 years of clinical imaging experience will record the presence and location of any abnormality, including gliosis, encephalomalacia, ischemia, volume loss, hydrocephalus, hemorrhage and/or malformations. Quantitative analysis will be performed using the volumetric 3D T1-weighted sequence using Freesurfer software (Version 5.3.0) to calculate whole brain grey matter and white matter volumes. Segmentation of the brain will allow for determination of regional volumes and measurement of cortical thickness. Based on this analysis, children will be assigned a Magnetic Resonance Imaging classification system (MRICS) for children with CP categorical score that can be used for subpopulation analysis in exploratory Aim 3.^76^ **Table 4** highlights the 5 MRICS categories and the MRICS breakdown of all current children with ITB who would qualify for this study (n=91), the majority of which fall into categories A-C.^77^ Using the data acquired above, Kralik and Thomas will manually map areas with abnormal anatomy onto known pediatric functional connectivity maps, likely the ABCD1000 and GSP1000.^78,79^ Descriptive analyses of variations and association with other CNS measures will be generated. Finally, each child’s DTI data will be analyzed using TBSS using FSL software which allows for whole brain quantitative analysis of the white matter tracts fractional anisotropy (FA), radial diffusivity (RD), axial diffusivity (AD), and mean diffusivity (MD).^80^

**Table 4.** MRICS categories and current ITB program distribution. This table shows the 5 MRICS categories on the left as well as the distribution of current children in our ITB program who would be eligible for this study.

| IMAGING FINDING | NUMBER | PERCENTAGE |
| --- | --- | --- |
| A. Maldevelopments | 16 | 18% |
| B. Predominant white matter injury | 28 | 31% |
| C. Predominant grey matter injury | 44 | 48% |
| D. Miscellaneous | 2 | 2% |
| E. Normal | 1 | 1% |

#### Missing Data

We have made a conservative assumption that 15% may drop out by 12-months. Even with best efforts to minimize missing data, we recognize some assessments will be missed or only partially completed. After systematically analyzing patterns of missing values, random multiple regression imputation will be used to assess the sensitivity of our conclusions to missing data. Baseline values will be imputed based on other baseline variables. Follow-up outcomes will be imputed from the previously observed outcomes. Analyses based on completed datasets will follow Rubin’s rules for combining multiple-imputed datasets.^81^ Multiple imputation methods are guided by the assumption that data are missing at random; subsequent analyses will also consider models for longitudinal changes, informed by findings about dropout patterns (if these appear to be strongly non-random and need other statistical adjustments). ^82,83^

## ETHICS AND DISSEMINATION

This study has been approved by the Baylor College of Medicine Institutional Review Board, 8/7/2024-6/25/2025. Protocol number H-54449. Title: “Intrathecal baclofen and pediatric dystonia: a clinical trial.” All research in this study will be conducted under the approval and monitoring of both an institutional review board as well as a data and safety monitoring committee. All parents of patients will be consented for this study. Parents and patients have the option to withdraw from the study at any time. All HIPPA will be protected. All data will be stored on encrypted, password-secured computers, and in encrypted, password protected servers, such as Redcap. Any printed files will be stored in the locked offices of the PI, research assistant, and data entry assistant. Any unnecessary files or duplicates will be permanently destroyed. Parents will be provided a written IRB-approved consent form. Written forms are currently available in English and Spanish (translation completed by TCH Language Services).

## PATIENT AND PUBLIC INVOLVEMENT

This study is motivated by the Research CP: Dystonia Edition, a clinician, researcher, and community member-driven Delphi study that was completed to identify research priorities in the area.^84^ The number 3 research theme was comparing the effectiveness of pharmacological and surgical treatments for dystonia (including evaluation of side effects, a person’s overall function, and effect on individualized goals). Our investigator team will engage parents and patients throughout the study to increase the awareness of and sensitivity to their concerns and observations with a formal Parent/Patient Council. The goal is to improve the relevance of research, patient safety, patient comfort, and overall study recruitment and cohort maintenance.^85,86^ We will recruit members from the TCH ITB program or past participants. We will aim to recruit at least one father and one adolescent/young adult with CP, ideally with personal ITB experience. We will plan for 2 virtual meetings annually. The Council will assist with this study in 1) reviewing appropriateness and clarity of recruitment methods and materials, 2) informed consent process, 3) clarity of the explanation of study assessment measures, and 4) plans for disseminating study results.

## CONCLUSION

The resulting rich dataset will improve patient selection for ITB by identifying key patient characteristics associated with positive outcomes and greatly inform future studies by identifying essential measures to efficiently assess clinical outcomes. Our proposed composite measure could serve as a superior outcome measure for future work on dystonic CP, a population in pressing need of efficacious treatment.

## Data Availability

All data produced in the present study are available upon reasonable request to the authors.

## AUTHOR CONTRIBUTIONS

ST, DF, SR conceptualized the study. ST, DF, SR, MC, SK, JR assisted in the design of the study. ST, DF, SR, MC, SK, JR funding procurement. ST wrote the initial draft of the manuscript and ST, DF, SR, MC, SK, JR revised the manuscript and approved of the final submission.

## REFERENCES

1. Centers for Disease Control and Prevention. Economic costs associated with mental retardation, cerebral palsy, hearing loss, and vision impairment. MMWR. Morbidity and mortality weekly report 2004;53:57–59.

2. Rice J, Skuza P, Baker F, et al. Identification and measurement of dystonia in cerebral palsy. Developmental medicine and child neurology 2017;59(12):1249–1255.

3. Parkes J, Dolk H HN. Children and young people with cerebral palsy in Northern Ireland - birth years 1977-1997. A comprehensive report from the Northern Ireland Cerebral Palsy Register. Belfast: 2005.

4. Himmelmann K, Hagberg G, Wiklund LM, et al. Dyskinetic cerebral palsy: a population-based study of children born between 1991 and 1998. Developmental medicine and child neurology 2007;49(4):246–51.

5. Reid SM, Meehan EM, Reddihough DS, Harvey AR. Dyskinetic vs Spastic Cerebral Palsy: A Cross-sectional Study Comparing Functional Profiles, Comorbidities, and Brain Imaging Patterns. Journal of child neurology 2018;33(9):593–600.

6. Bohn E, Goren K, Switzer L, et al. Pharmacological and neurosurgical interventions for individuals with cerebral palsy and dystonia: a systematic review update and meta-analysis. Developmental Medicine and Child Neurology 2021;63(9):1038–1050.

7. Sanger TD, Delgado MR, Gaebler-Spira D, et al. Classification and definition of disorders causing hypertonia in childhood. Pediatrics 2003;111(1):e89–97.

8. Graham D, Paget SP, Wimalasundera N. Current thinking in the health care management of children with cerebral palsy. The Medical journal of Australia 2019;210(3):129–135.

9. Harvey A, Reddihough D, Scheinberg A, Williams K. Oral medication prescription practices of tertiary-based specialists for dystonia in children with cerebral palsy. Journal of paediatrics and child health 2018;54(4):401–404.

10. Penner M, Xie WY, Binepal N, et al. Characteristics of pain in children and youth with cerebral palsy. Pediatrics 2013;132(2):e407–13.

11. Monbaliu E, De Cock P, Mailleux L, et al. The relationship of dystonia and choreoathetosis with activity, participation and quality of life in children and youth with dyskinetic cerebral palsy. European journal of paediatric neurology : EJPN : official journal of the European Paediatric Neurology Society 2017;21(2):327–335.

12. Rice J, Russo R, Halbert J, et al. Motor function in 5-year-old children with cerebral palsy in the South Australian population. Developmental medicine and child neurology 2009;51(7):551–6.

13. Albright AL, Ferson SS. Intrathecal baclofen therapy in children. Neurosurgical focus 2006;21(2):e3.

14. Hasnat MJ, Rice JE. Intrathecal baclofen for treating spasticity in children with cerebral palsy. The Cochrane database of systematic reviews 2015;(11):CD004552.

15. Albright AL. Baclofen in the treatment of cerebral palsy. Journal of Child Neurology 1996;11(2):77–83.

16. Gracies JM, Nance P, Elvoic E, McGuire J SD. Traditional pharmacological treatments for spasticity. Part II: General and regional treatments. Muscle Nerve Suppl 1997;6:S92–120.

17. Malcangio M BN. Gamma-aminobutyric acidB, but not gamma-aminobutyric acidA receptor activation, inhibits electrically evoked substance P-like immunoreactivity release from the rat spinal cord in vitro. Journal of Pharmacological and Experimental Therapeutics 1993;266(3):1490–1496.

18. Narayan RK, Loubser PG, Jankovic J, et al. Intrathecal baclofen for intractable axial dystonia. Neurology 1991;41(7):1141–2.

19. Albright AL, Barry MJ, Painter MJ, Shultz B. Infusion of intrathecal baclofen for generalized dystonia in cerebral palsy. Journal of neurosurgery 1998;88(1):73–6.

20. Dvorak EM, McGuire JR, Nelson MES. Incidence and identification of intrathecal baclofen catheter malfunction. PM & R : the journal of injury, function, and rehabilitation 2010;2(8):751–6.

21. Spader HS, Bollo RJ, Bowers CA, Riva-Cambrin J. Risk factors for baclofen pump infection in children: a multivariate analysis. Journal of neurosurgery. Pediatrics 2016;17(6):756– 62.

22. Feller CN, Awad AJ, Nelson MES, et al. Low Rate of Intrathecal Baclofen Pump Catheter-Related Complications: Long-Term Study in Over 100 Adult Patients Associated With Reinforced Catheter. Neuromodulation : journal of the International Neuromodulation Society 2021;24(7):1176–1180.

23. de Lissovoy G, Matza LS, Green H, et al. Cost-effectiveness of intrathecal baclofen therapy for the treatment of severe spasticity associated with cerebral palsy. Journal of child neurology 2007;22(1):49–59.

24. Albright AL, Barry MJ, Shafton DH, Ferson SS. Intrathecal baclofen for generalized dystonia. Developmental medicine and child neurology 2001;43(10):652–7.

25. Motta F, Stignani C, Antonello CE. Effect of intrathecal baclofen on dystonia in children with cerebral palsy and the use of functional scales. Journal of Pediatric Orthopaedics 2008;28(2):213–217.

26. Motta F, Antonello CE, Stignani C. Upper limbs function after intrathecal baclofen therapy in children with secondary dystonia. Journal of pediatric orthopedics [date unknown];29(7):817–21.

27. Motta F, Antonello CE SC. Intrathecal baclofen and motor function in cerebral palsy. Dev Med Child Neurol 2011;53:443–448.

28. Eek MN, Olsson K, Lindh K, et al. Intrathecal baclofen in dyskinetic cerebral palsy: effects on function and activity. Developmental medicine and child neurology 2018;60(1):94–99.

29. Kim JH, Jung NY, Chang WS, et al. Intrathecal Baclofen Pump Versus Globus Pallidus Interna Deep Brain Stimulation in Adult Patients with Severe Cerebral Palsy. World neurosurgery 2019;126:e550–e556.

30. Bonouvrié LA, Becher JG, Vles JSH, et al. The Effect of Intrathecal Baclofen in Dyskinetic Cerebral Palsy: The IDYS Trial. Annals of Neurology 2019;86(1):79–90.

31. Bonouvrié LA, Haberfehlner H, Becher JG, et al. Attainment of personal goals in the first year of intrathecal baclofen treatment in dyskinetic cerebral palsy: a prospective cohort study. Disability and rehabilitation 2022;1–8.

32. Bonouvrie LA, Haberfehlner H, Becher JG, Vles JSH, Vermeulen J BA. Attainment of personal goals in the first year of intrathecal baclofen treatment in dyskinetic cerebral palsy: a prospective cohort study. Disability and Rehabilitation 2023;45(8):1315–1322.

33. van de Pol LA, Vermeulen RJ, van ’t Westende C, et al. Erratum: Risk Factors for Dystonia after Selective Dorsal Rhizotomy in Nonwalking Children and Adolescents with Bilateral Spasticity. Neuropediatrics 2018;49(1):e1.

34. Gober J, Seymour M, Miao H, et al. Management of severe spasticity with and without dystonia with intrathecal baclofen in the pediatric population: a cross-sectional study. World Journal of Pediatric Surgery 2022;

35. Stephen CD, Simonyan K, Ozelius L, Breakefield XO SN. Chapter 40 - Dystonia. In: Zigmond MJ, Wiley CA CM-F, editor. Neurobiology of Brain Disorders. Academic Press; 2023 p. 713–751.

36. Kuyper DJ, Parra V, Aerts S, Okun MS KB. Nonmotor manifestations of dystonia: a systematic review. Mov Disord 2011;26(7):1206–1217.

37. Graefe SB MS. Substance P. In: StatPearls. Treasure Island, FL: StatPearls Publishing; 2022

38. Jinnah HA, Neychev V, Hess EJ. The Anatomical Basis for Dystonia: The Motor Network Model. Tremor and other hyperkinetic movements (New York, N.Y.) 2017;7:506.

39. Papadimitriou I, Dalivigka Z, Outsika C, et al. Dystonia assessment in children with cerebral palsy and periventricular leukomalacia. European journal of paediatric neurology : EJPN : official journal of the European Paediatric Neurology Society 2021;32:8–15.

40. Drougia A, Giapros V, Krallis N, et al. Incidence and risk factors for cerebral palsy in infants with perinatal problems: A 15-year review. Early Human Development 2007;83(8):541–547.

41. Aravamuthan BR, Waugh JL. Localization of Basal Ganglia and Thalamic Damage in Dyskinetic Cerebral Palsy. Pediatric neurology 2016;54:11–21.

42. Park B-H, Park S-H, Seo J-H, et al. Neuroradiological and neurophysiological characteristics of patients with dyskinetic cerebral palsy. Annals of rehabilitation medicine 2014;38(2):189–99.

43. Arens LJ, Peacock WJ, Peter J. Selective posterior rhizotomy: a long-term follow-up study. Child’s nervous system : ChNS : official journal of the International Society for Pediatric Neurosurgery 1989;5(3):148–52.

44. Barry MJ, VanSwearingen JM, Albright AL. Reliability and responsiveness of the Barry-Albright Dystonia Scale. [Internet]. Dev Med Child Neurol 1999;41(6):404–11.Available from: http://www.ncbi.nlm.nih.gov/pubmed/10400175

45. Jethwa A, Mink J, Macarthur C, et al. Development of the Hypertonia Assessment Tool (HAT): a discriminative tool for hypertonia in children [Internet]. Dev Med Child Neurol 2010;52(5)Available from: https://onlinelibrary.wiley.com/doi/10.1111/j.1469-8749.2009.03483.x

46. F M, C S, CE. A. Effect of intrathecal baclofen on dystonia in children with cerebral palsy and the use of functional scales. J Pediatr Orthop 2008; [date unknown];28:213–217.

47. F M, C S, CE. A. The use of intrathecal baclofen pump implants in children and adolescents: safety and complications in 200 consecutive cases. J Neurosurg 2007; 107(S1): 32–5; [date unknown].

48. Leland Albright A, Barry MJ, Shafron DH, Ferson SS. Intrathecal baclofen for generalized dystonia. Dev Med Child Neurol 2001;43(10):652–657.

49. Bonouvrié LA, Becher JG, Vles JSH, et al. Intrathecal baclofen treatment in dystonic cerebral palsy: A randomized clinical trial: The IDYS trial. BMC Pediatr 2013;13(1):1–8.

50. Boster AL, Adair RL, Gooch JL, et al. Best Practices for Intrathecal Baclofen Therapy: Dosing and Long-Term Management. Neuromodulation 2016;19(6):623–631.

51. Stewart K, Lewis J, Wallen M, et al. The Dyskinetic Cerebral Palsy Functional Impact Scale: development and validation of a new tool [Internet]. Dev Med Child Neurol 2021;63(12):1469–1475.Available from: https://onlinelibrary.wiley.com/doi/10.1111/dmcn.14960

52. Barry MJ, VanSwearingen JM, Albright AL. Reliability and responsiveness of the Barry-Albright Dystonia Scale. Developmental medicine and child neurology 1999;41(6):404–11.

53. Monbaliu E, Ortibus E, Roelens F, Desloovere K, Deklerck J, Prinzie P, de Cock P FH. Rating scales for dystonia in cerebral palsy: reliability and validity. Dev Med Child Neurol 2010;52:570–575.

54. Bollo RJ, Gooch JL WM. Stereotactic endoscopic placement of third ventricle catheter for long-term infusion of baclofen in child with secondary generalized dystonia. J Neurosurg Pediatr 2012;10:30–33.

55. Ward A, Hayden S, Dexter M, Scheinberg A. Continuous intrathecal baclofen for children with spasticity and/or dystonia: Goal attainment and complications associated with treatment. Journal of paediatrics and child health 2009;45(12):720–6.

56. Stewart K, Harvey A JL. A systematic review of scales to measure dystonia and choreoathetosis in children with dyskinetic cerebral palsy. Dev Med Child Neurol 2017;59(8):785–795.

57. Vanmechelen I, Danielsson A, Lidbeck C, et al. The Dyskinesia Impairment Scale, Second Edition: Development, construct validity, and reliability. Developmental Medicine & Child Neurology 2023;65(5):683–690.

58. Monbaliu E, Orbitus E, De Cat, J, Dan B, Heyrman L, Prinzie P, De Cock P FH. The Dyskinesia Impairment Scale: a new instrument to measure dystonia and choreoathetosis in dyskinetic cerebral palsy. Dev Med Child Neurol 2012;54:278–283.

59. Teixeira AL, Maia DP CF. UFMG Sydenham’s chorea rating scale (USCRS): reliability and consistency. Movement Disorders 2005;20:585–591.

60. Grisold W. The expanding burden of neurological disorders. Lancet Neurol 2024;23(4):326–327.

61. Love S, Gibson N, Smith N, et al. Interobserver reliability of the Australian Spasticity Assessment Scale (ASAS). Developmental medicine and child neurology 2016;58 Suppl 2:18–24.

62. Mutlu A, Livanelioglu A GM. Reliability of Ashworth and Modified Ashworth Scales in children with spastic cerebral palsy. BMC Musculoskel Dis 2008;9:44.

63. Bjornson K, Graubert C MJ. Test-retest reliability of the gross motor function measure in children with cerebral palsy. Pediatric Physical Therapy 2000;12(4):200–202.

64. Mahasup N, Sritipsukho P, Lekskulchai R KP. Inter-rater and intra-rater reliability of the gross motor function measure (GMFM-66) by Thai pediatric physical therapists. Journal of the Medical Association of Thailand 2011;94(S7):S139–S144.

65. Oeffinger D, Bagley A, Rogers S, Gorton G, Kryscio R, Abel M, Damiano D, Barnes D TC. Outcome tools used for ambulatory children with cerebral palsy: responsiveness and minimally clinically important differences. Dev Med Child Neurol 2008;50(12):918–925.

66. Platz T, Pinkowski C, van Wijck F, K I-H, di Bella P JG. Reliability and validity of arm function assessment with standardized guidelines for the Fugl-Meyer Test, Action Research Arm Test and Box and Block Test: a multicentre study. Clin Rehabil 2005;19(4):404–411.

67. Liang KJ, Chen HL, Shieh JY WT-N. Measurement properties of the box and block test in children with unilateral cerebral palsy. Science Reports 2021;11:20955.

68. Martini R, Rios J, Polatajko H, et al. The performance quality rating scale (PQRS): reliability, convergent validity, and internal responsiveness for two scoring systems. Disability and rehabilitation 2015;37(3):231–8.

69. Cusick A, Lannin NA, Lowe K. Adapting the Canadian Occupational Performance Measure for use in a paediatric clinical trial. Disability and Rehabilitation 2007;29(10):761–766.

70. Narayanan UG, Fehlings D, Weir S, Knights S, Kiran S CK. Initial development and validation of the Caregiver Priorities and Child Health Index of Life with Disabilities (CPCHILD). Dev Med Child Neurol 2006;48:804–812.

71. Steenbeek D, Gorter JW, Ketelaar M, Galama K LE. Responsiveness of Goal Attainment Scaling in comparison to two standardized measures in outcome evaluation of children with cerebral palsy. Clin Rehabil 2011;25(12):1128–1139.

72. Johnson R, Wichern D. Applied multivariate statistical analyses. 6th ed. London: Pearson Classic Series; 2007.

73. Tibshirani R, Walther G, Hastie T. Estimating the number of clusters in a dataset via the gap statistic. Journal of the Royal Statistical Society, Ser B. 2001;63(2):411–423.

74. Johnson R, Wichern D. Applied multivariate statistical analyses. 6th ed. London: Pearson Classic Series; 2007.

75. Burns TM, Conaway M, Sanders DB. The MG Composite. Neurology 2010;74(18):1434– 1440.

76. Himmelmann K, Horber V, De La Cruz J, et al. MRI classification system (MRICS) for children with cerebral palsy: development, reliability, and recommendations. Developmental Medicine & Child Neurology 2017;59(1):57–64.

77. Joutsa J, Corp DT, Fox MD. Lesion network mapping for symptom localization: recent developments and future directions. Current Opinion in Neurology 2022;35(4):453–459.

78. Fox MD. Mapping Symptoms to Brain Networks with the Human Connectome. New England Journal of Medicine 2018;379(23):2237–2245.

79. Lumsden DE, Ashmore J, Ball G, et al. Fractional anisotropy in children with dystonia or spasticity correlates with the selection for DBS or ITB movement disorder surgery. Neuroradiology 2016;58(4):401–8.

80. Rubin D. Multiple imputation for nonresponse in surveys. New York, NY: John Wiley and Sons; 1987.

81. Su L, Li Q, Barrett JK, Daniels MJ. A sensitivity analysis approach for informative dropout using shared parameter models. Biometrics 2019;75(3):917–926.

82. Diggle P, Kenward MG. Informative Drop-Out in Longitudinal Data Analysis. Applied Statistics 1994;43(1):49.

83. Gilbert LA, Fehlings DL, Gross P, et al. Top 10 Research Themes for Dystonia in Cerebral Palsy [Internet]. Neurology 2022;99(6):237–245.Available from: http://www.neurology.org/lookup/doi/10.1212/WNL.0000000000200911

84. Roennow A, Sauvé M, Welling J, et al. Collaboration between patient organisations and a clinical research sponsor in a rare disease condition: learnings from a community advisory board and best practice for future collaborations. BMJ open 2020;10(12):e039473.

85. Brockman TA, Balls-Berry JE, West IW, et al. Researchers’ experiences working with community advisory boards: How community member feedback impacted the research. Journal of clinical and translational science 2021;5(1):e117.

